# Social multipliers and the Covid-19 epidemic: Analysis through constrained maximum entropy modeling

**DOI:** 10.1101/2020.09.01.20185868

**Authors:** Duncan K. Foley

## Abstract

Social multipliers occur when individuals’ actions influence other individuals’ actions so as to lead to amplified aggregate effects. Epidemic infections offer a dramatic example of this phenomenon since individual actions such as social distancing and masking that have small effects on individual risk can have very large effects in reducing rates of infection when they are widely adopted. This paper uses the info-metric method of constrained maximum entropy modeling to estimate the impact of social multiplier effects in the Covid-19 epidemic with a model that infers the length of infection, the rate of mortality, the base infection factor, and reductions in the infection factor due to changes in social behavior from data on daily infections and deaths. When the model takes account of the rate of reporting of infections, it produces two scenarios of epidemic dynamics: one in which reporting is low, under 10%, the estimated infection is correspondingly large, and immunity effects play a significant role in stabilizing the epidemic; and a second where reporting rates are close to 100%, and the epidemic is controlled mostly by changes in social behavior.

## Introduction

### Social interaction and epidemics

The phenomenon of social multipliers, social interactions in which the impact of individual behavioral decisions have amplified social effects due to their impact on other members of society, are a pervasive feature of economic and social life. Social multiplier effects dominate important social-economic phenomena such as the determination of aggregate demand and employment in capitalist economies, asset prices in speculative markets, financial and economic crises, revolutionary political movements, and traffic congestion. In viral epidemics the social multiplier effect appears because individuals’ attempts to protect themselves against infection through social isolation, reduced social contact, and masking have a multiplied impact on the social spread of infection by changing the probability of that an infectious individual will infect others.

### Covid-19 and social multipliers

The case of Covid-19 is particularly instructive in this respect, because the mean length of the infectious period of the disease is relatively short, on the order of 5-10 days, but it is so infectious that its spread through aerosols and other mechanisms in public spaces can double the number of infected also in a matter of days. Under these circumstances even small changes in behavior will have greatly amplified effects on the spread an epidemic on a short time horizon through the social multiplier. Each individual can do very little to protect themselves from a widespread and rapidly growing epidemic, but the collective effect of changes in social behavior that reduce the probability of infections can rapidly bring the epidemic under control.

### A new method of empirical analysis

This paper presents a method for the empirical analysis of epidemic statistics consisting of daily reports of new infections and deaths, and applies the model to data on national Covid-19 epidemics from the European Center for Disease Prevention and Control database. The method is the constrained maximum entropy framework described by [2]. In the context of epidemic modeling, this framework provides a method for estimating the unobserved daily rate of infection together with estimates of the fraction of infections reported as “confirmed”, and average recovery and mortality rates from daily reports of new confirmed infections and deaths.

### Summary of results

The results of this study identify the expected length of Covid-19 infectiousness as on the order of 5-10 days, and the base infection factor as on the order of two new infections per existing infection. The qualitative pattern of infection dynamics is quite sensitive to the estimate of the reporting rate, which is only weakly identified by the model using this limited data. If the reporting rate is constrained to be close to 100%, the model estimates small epidemics with high death rates which are brought under control primarily by changes in social behavior. If the reporting rate is estimated without constraint, the model for many cases estimates a reporting rate under 10%, resulting in a very large estimated epidemic with a lower death rate, which is brought under control in part by the reduction in the susceptible population.

## Epidemic modeling SIR Model

### SIR (SFR) Model

On a given day of an epidemic *t* = 1, …, *T*, let *S*_*t*_ ≥ 0 be the proportion of a population uninfected and therefore susceptible, *F*_*t*_, ≥ 0 the proportion of the population infected and infectious, and *R*_*t*_ = 1 − *S*_*t*_, − *F*_*t*_ the proportion of the population either recovered or died, and therefore unsusceptible. *T* is the number of days over which the model tracks the epidemic, starting from day 1. We write *F* rather than *I* for the infected population.

### SIR Equations

These variables follow the deterministic laws:

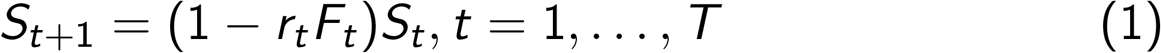

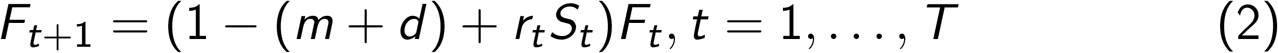

Here *r*_*t*_ is the infection rate for an encounter between a susceptible and infected person given the pattern of social behavior at day *t*, and *m* and *d* are the recovery and death rates for infections, which apply uniformly to the current infected population. In this context *m* is the rate at which an infected individual stops being infectious, not necessarily the rate at which infected individuals become symptom-free. This is a version of the well-known SIR dynamic model of epidemics [3]. (For a discussion of some of the limits of the SIR model, see [1].)

This type of geometric recovery and death rate implies that the expected length of an infection is 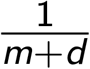.

### Epidemiological features

This model assumes that recoveries and deaths occur uniformly among the stock of currently infected individuals, which is at best an approximation, since we know the likelihood of recovery or death in fact depend on the length of the infection. Given the limitations of the data we have to work with, this approximation seems unavoidable, and the model fits reported below indicate that it does not compromise the ability of the model to track epidemics.

### Dynamic analysis

This stripped down SIR model is a two-dimensional discrete-time nonlinear dynamical system. The linear approximation to this dynamical system is:

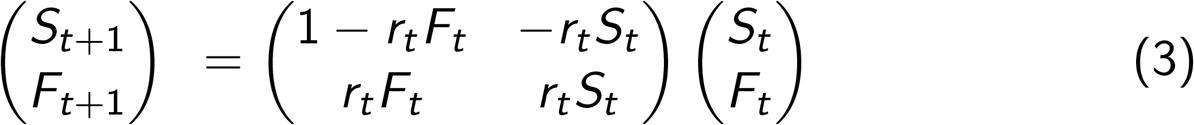

The eigenvalues of this system at *F*_*t*_ = 0 are inside the unit circle, and therefore stable, if *r*_*t*_*S*_*t*_ *< m* + *d* or 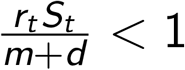. When this condition is met, epidemics die out because the reproduction factor for infections is less than unity. The model parameters *r*_*t*_ are the daily rate of infections, and to translate them into the more frequently used infection factor it is necessary to multiply the daily infection rate by the expected length of an infection in days, 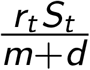.

### Constrained maximum entropy (CME) modeling

Constrained maximum entropy (CME) modeling represents the theoretical principles defining the system that produced the data as constraints. These constraints restrict the set of joint probability distributions over the states of the system, but typically the constraints are not sufficient to determine the joint probability distribution. The resulting underdetermination is formally resolved by ranking the feasible joint probability distributions according to their Shannon information entropy. The feasible joint probability distribution with the highest informational entropy is the CME estimate of the system’s unobserved states. Ranking probability distributions by their informational entropy ensures that all the information shaping the result enters through the explicit constraints.

### CME for the SIR model

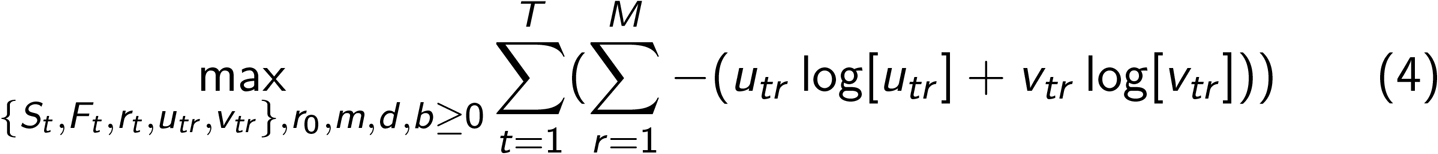

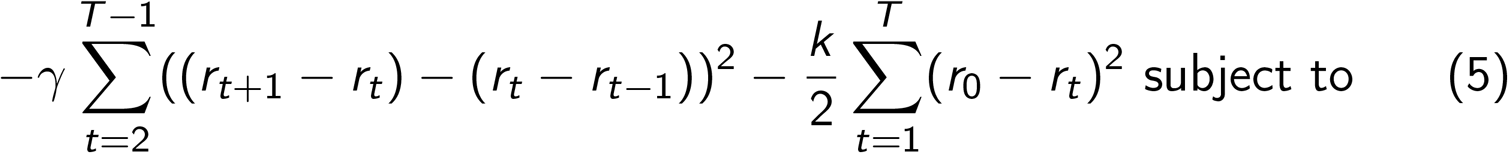

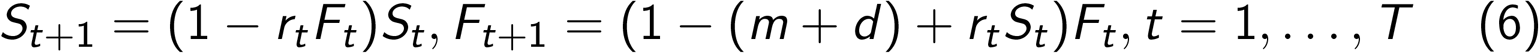

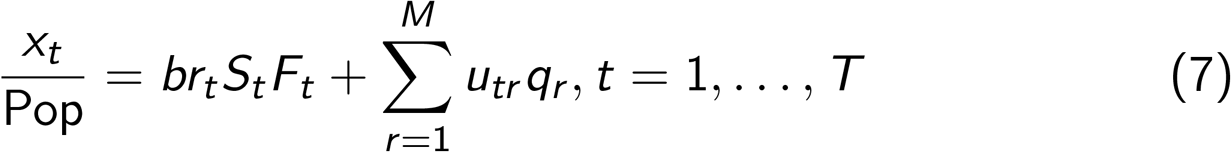

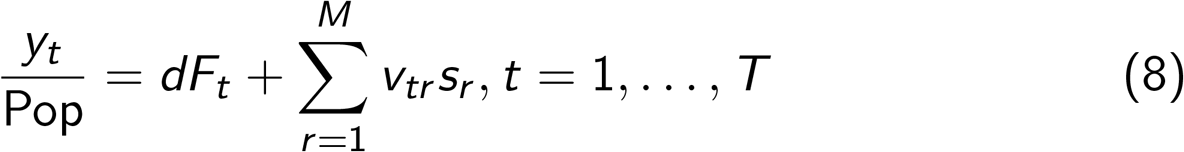

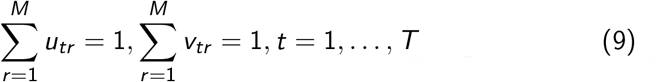

### Details of CME Setup – Error distributions

Here *u*_*tr*_, *v*_*tr*_ are error frequency distributions over the *M*-dimensional supports *q*_*r*_, *s*_*r*_, the expectations of which constitute the errors in fitting the observed daily data on infections, *x*_*t*_, and mortality, *y*_*t*_, given the fraction of actual infections reported as “confirmed”, *b*. The supports are chosen symmetrically to include three (this factor appears in the documentation as the “supportwidth” parameter) standard deviations of the corresponding data series, so that the maximum entropy of the frequency distributions corresponds to zero errors, or, equivalently, errors require a reduction in the entropy of the frequency distributions. The maximization of entropy of the error frequency distributions implies minimization of the errors.

### Details of CME – Smoothing

The objective function has two other terms.

The first penalizes the variation in *r*_*t*_ represented by the square of second differences. The weight on this factor in the simulations below is *γ* = 1000, which results in smoothing daily fluctuations in the estimated infection rates. Experimentation indicates that the qualitative results of the simulations are robust to variations in this parameter.

### Details of CME – Resistance

The second additional term penalizes the deviation of the daily infection rate, *r*_*t*_ below the base infection rate, *r*_0_. The idea is that changing social behavior to depress *r*_*t*_ below the base level *r*_0_ requires social “force” similar to the physical force depressing a spring, and results in a potential energy 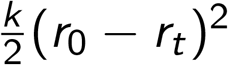, which trades off against the entropy of the frequency distributions of error terms on the observed data. This off-setting force we call *resistance*. The reduction in the daily infection rate is partly a spontaneous reaction of individuals to their recognition that an epidemic is spreading by reducing their exposure, and partly a social response mediated by legal guidelines and the development of social norms concerning masking, social distancing, and the like. Since these changes in behavior have substantial economic and other costs, it makes sense to include a parameter such as *k* to represent those costs. In the simulations reported below typically 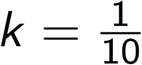, a value that is in a range that produces enough resistance to changing social behavior to require a reasonable level for the infection factor 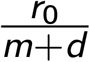.

### Identifying base infection level

The inclusion of this term is also necessary to allow the model to identify the base infection level, *r*_0_. The only evidence available on which to make an estimate of this base infection rate is the growth of new infections in the first few days of the epidemic, before changes in social behavior have begun to affect *r*_*t*_. But the number of new infections is an inherently noisy variable, and estimates of *r*_0_ through extrapolation of the number of new infections turns out to be highly unstable empirically. The inclusion of the resistance in the objective function has the effect of making the estimates of the daily infection rate in the days before the epidemic takes hold uniform over time, and of stabilizing these estimates.

### Logic of simulation

The logic of the simulation method is as follows. We have data on observed new infections, *x*_*t*_, and deaths, *y*_*t*_, for T days, and want to infer the history of the epidemic, the vectors {*S*_*t*_} and {*F*_*t*_}, the unobserved daily infection rates, {*r*_*t*_} and the parameters *r*_0_, *m, d, b*, by finding the values of these parameters that maximize the sum of the entropy of the error distributions and the penalty functions subject to the constraints (4).^2^

### Example: United States

**Figure 1:**
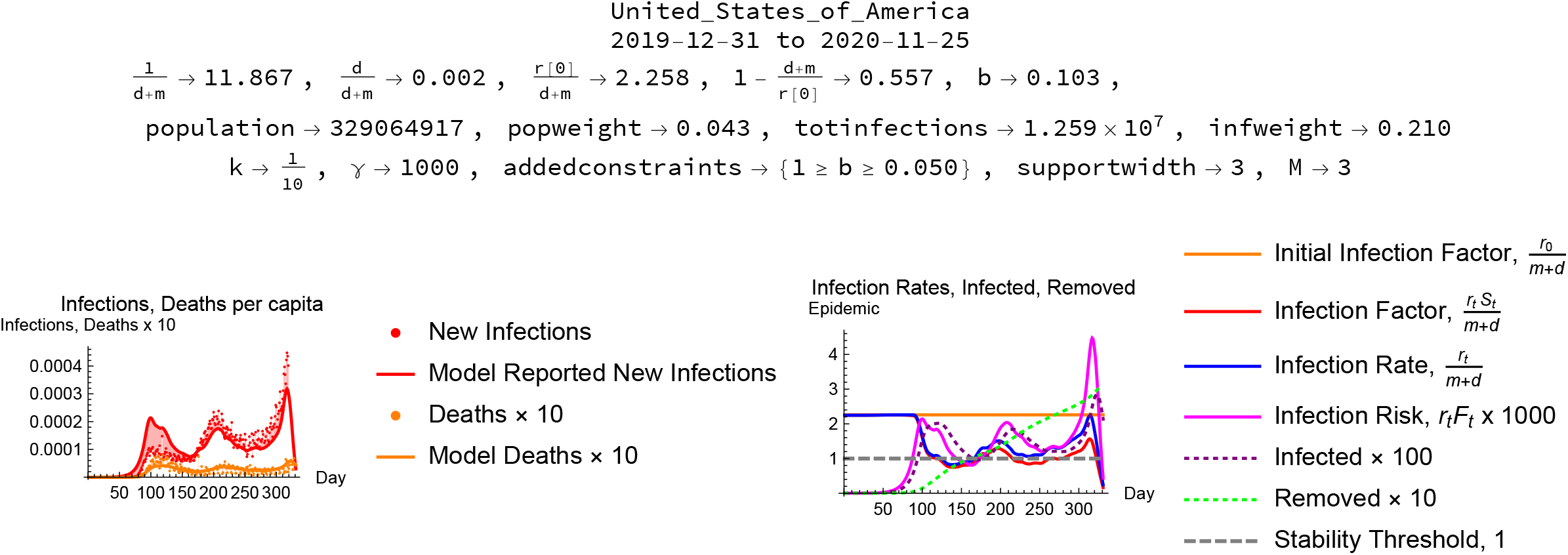
Simulation for the U.S.

### Reading the simulation – Parameter estimates

Figure 1 shows the results of the simulation for data from the United States. The heading documents the country and period of the simulation, reports the estimated mean length of the disease in days, 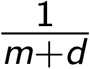, the estimated proportion of cases leading to death, 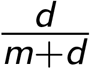, the estimated infection factor, 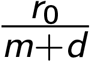, the implied threshold herd immunity (the proportion of the population that has to be immune to drive the infection factor below 1), 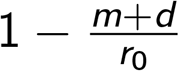, other background statistics, and the parameters used for the simulation. The model estimates the average duration of the infectious stage of the disease as about 12 days, the proportion of cases resulting in death as .2%, the infection factor without changes in social behavior about 2.5, the resulting herd immunity threshold as requiring 55% of the population to be immune to suppress the epidemic, and the reporting rate to be 10%.

### Reading the simulation – Plots

The left plot shows observed new infections per day, the model simulated new infections, infection errors, with the same statistics for deaths. The right plot shows the estimated initial infection factor, the estimated daily infection factor and infection rate, the rescaled daily infection risk, the infected population, and the population removed from susceptibility through recovery or death. Both plots have the days of the epidemic on the horizontal axis.

### Control of epidemics

An important feature of this simulation is that it indicates that the epidemic in the U.S. was controlled, to the extent it has been controlled, by both changes in social behavior that reduced the infection factor, and by herd immunity exhausting the pool of individuals susceptible to the infection. The red and blue curves in the lower-left plot dramatically record the fall in the infection rate a few days after the infection began to take hold, as well as a rebound in the infection rate after the first peak of the epidemic.

### Example: New York City

Data on infections and deaths from Covid-19 are also available for New York City, from https://www1.nyc.gov/site/doh/covid/covid-19-data.page, accessed November 27, 2020.

### Reading the NYC simulation

Figure 2 shows the results of the simulation for data from New York City with no constraint on the reporting rate. The model estimates the average duration of the infectious stage of the disease as about 7 days, the proportion of cases resulting in death as .6%, the infection factor without changes in social behavior about 1.9, and the resulting herd immunity threshold as requiring 47% of the population to be immune to suppress the epidemic. The reporting rate is estimated at about 7.5%, implying that for every “confirmed” infection there were about 13 unreported further infections. This simulation implies a very large epidemic eventually controlled by an approach to herd immunity with close to 50% of the population removed from the pool of susceptibles by recovery or death. (The resolution of the time series information on which this simulation is based makes the identification of re-infection as a significant factor problematic.) Retrospective serological tests for antibodies in New York City, by contrast, report at most about 20% of the population with antibodies indicating infection.

**Figure 2:**
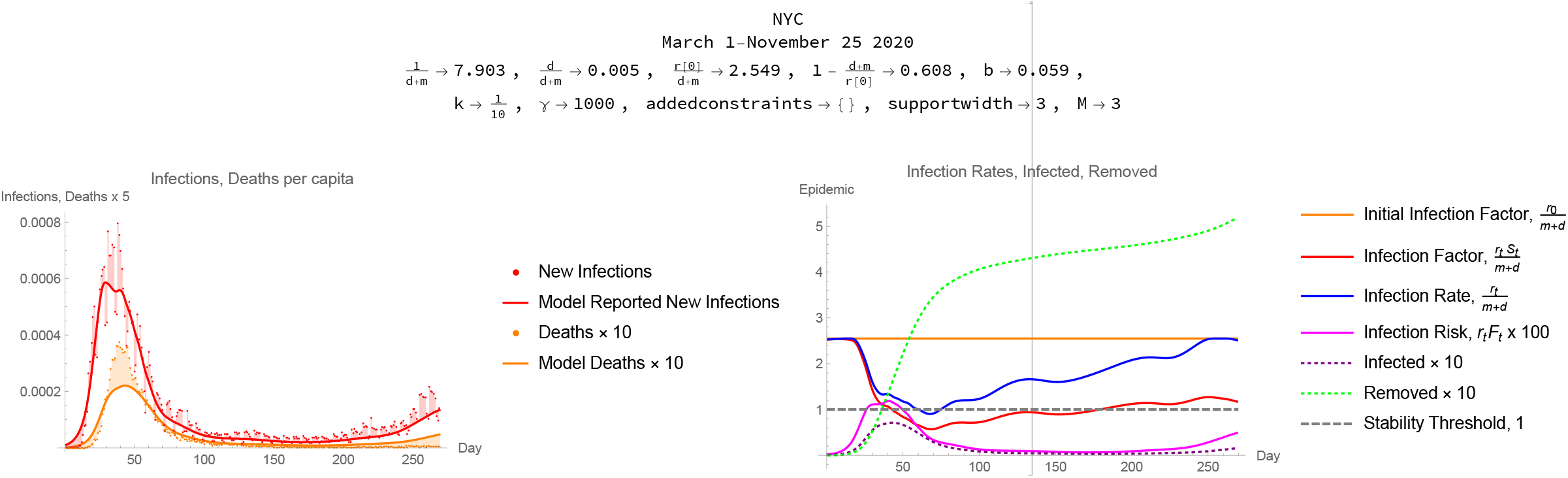
Simulation for New York City with no constraint on the reporting rate. The report has the same format as Figure <u>1.</u>.

### Limits of resolution

The uncertainty with which the method estimates the reporting rate and the size of the epidemic reflect limitations of the time series data we have as input. Although the CME method extracts all the information available from a data set and model, there is only so much information to be extracted. Further insight depends on adding further information in the form of constraints.

### Cross-country statistics

Data from the European Center for Disease Prevention and Control reports daily infection and mortality counts for 209 countries. The program reports errors or times out on some of these country data sets, but usable simulations result in countries. Some of these cases report small total numbers of infections. Summary statistics for the modeling of the 81 country epidemics reporting more than 10,000 infections are reported in Table 1.

**Table 1:** Infection-weighted median values of the epidemic parameters together

| Stat | Infection Weighted<br>Median | Interquartile Range |
| --- | --- | --- |
| $\frac{1}{d+m}$ | 11.867 | 0.381<br>16.200 |
| $\frac{d}{d+m}$ | 0.014 | 0.002<br>0.027 |
| $\frac{r[0]}{d+m}$ | 2.048 | 1.041<br>2.258 |
| $1 - \frac{d+m}{r[0]}$ | 0.512 | 0.039<br>0.557 |
| b | 1.000 | 0.103<br>1.000 |

$\{k \rightarrow \frac{1}{10}, \gamma \rightarrow 1000, \alpha \rightarrow 0, \text{addedconstraints} \rightarrow \{1 \geq b \geq 0.050\}, \text{supportwidth} \rightarrow 3, M \rightarrow 3\}$

### Noise

The parameter estimates, which in some cases depend on extrapolating early phases of epidemics, are predictably quite noisy, but the medians, 12 days for the expected infectious period, a 1.4% mortality rate among the infected, and a base infection factor 2, are within ranges frequently reported for these parameters.

### Country cases

Some national epidemics show basically the same pattern as the United States: an estimate of infection length between 4 and 12 days, an infection factor between 1.5 and 3, and a sharp drop in the daily infection factor presumably as a result of changes in social behavior such as isolation and masking motivated by a combination of individual protection and social norms backed up public policy. In many instances the model estimates that the infection factor rises again after a fall in infections, in some cases heralding a resurgence of the infection. Some national epidemics scenarios attribute a significant role in controlling epidemics to removal of a significant part of the population from the pool of susceptibles through recovery or death. These simulations all constrain the reporting rate to be between 10% and 100%, and in several cases the simulation maximizes the log fit at one or the other of these bounds. A selection of these cases appears here in Figures 3–8.

### China

**Figure 3:**
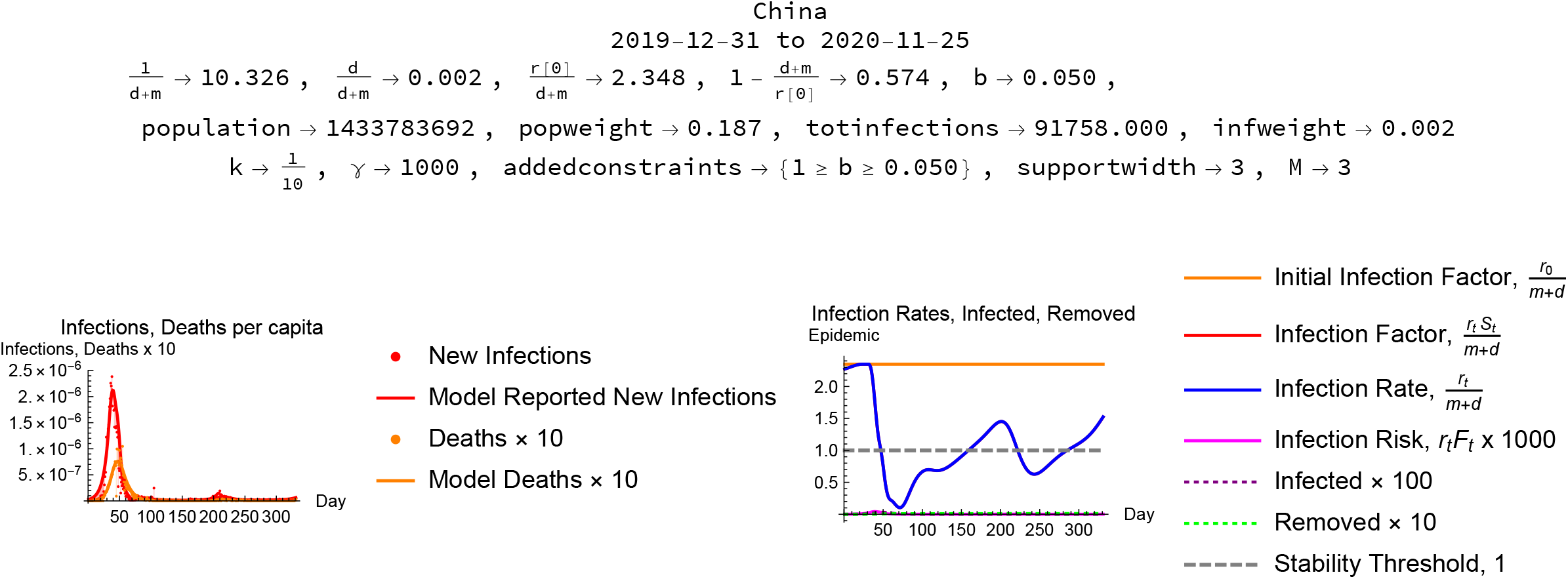
China. The sharp spikes in the infection and mortality data represent outliers that are excluded from the model simulation constraints.

### Italy

**Figure 4:**
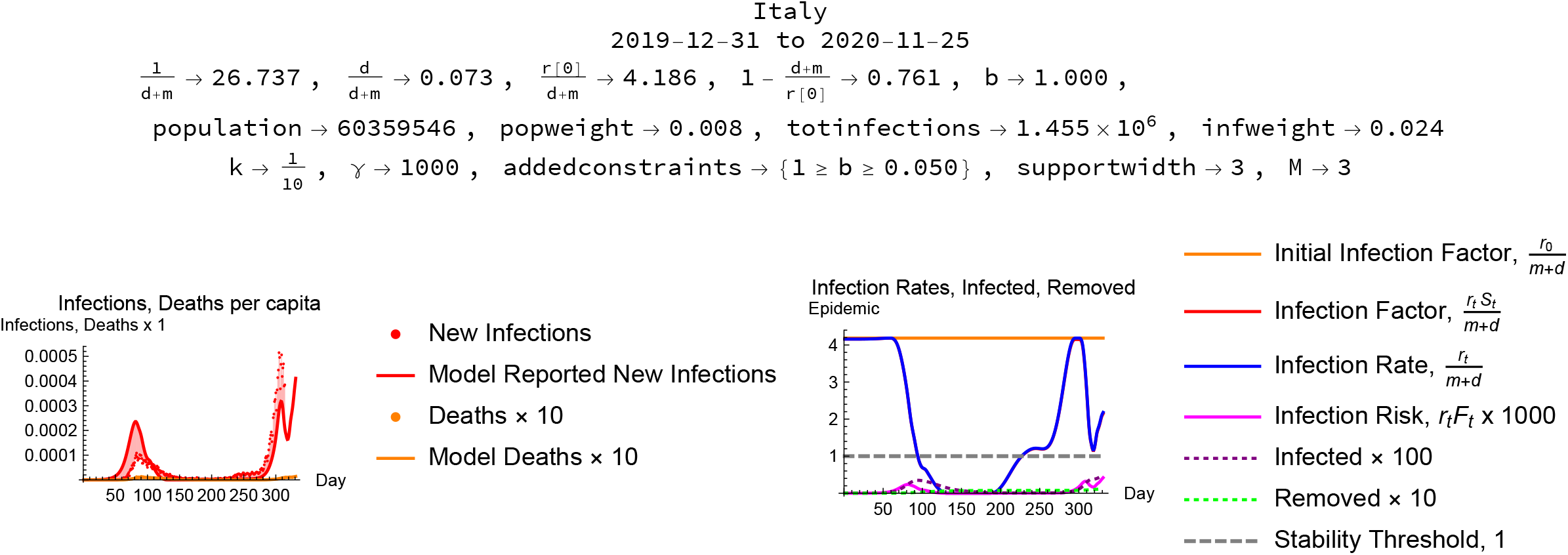
Italy.

### France

**Figure 5:**
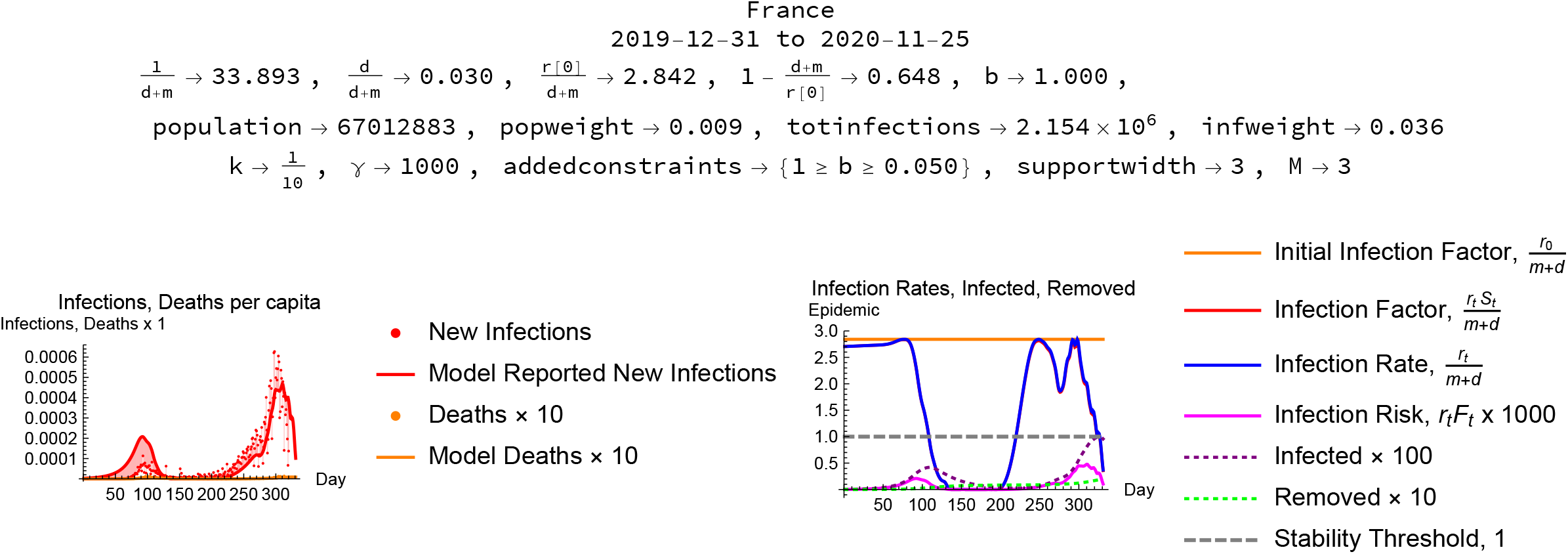
France.

### Germany

**Figure 6:**
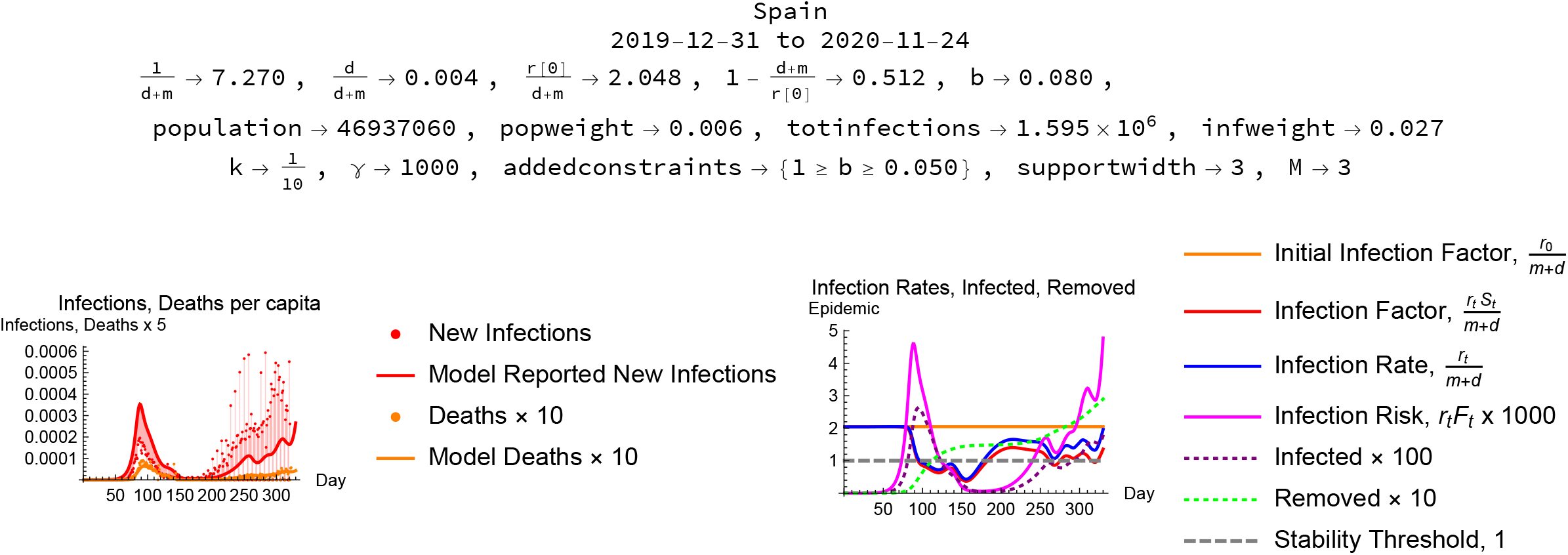
Spain.

### Russia

**Figure 7:**
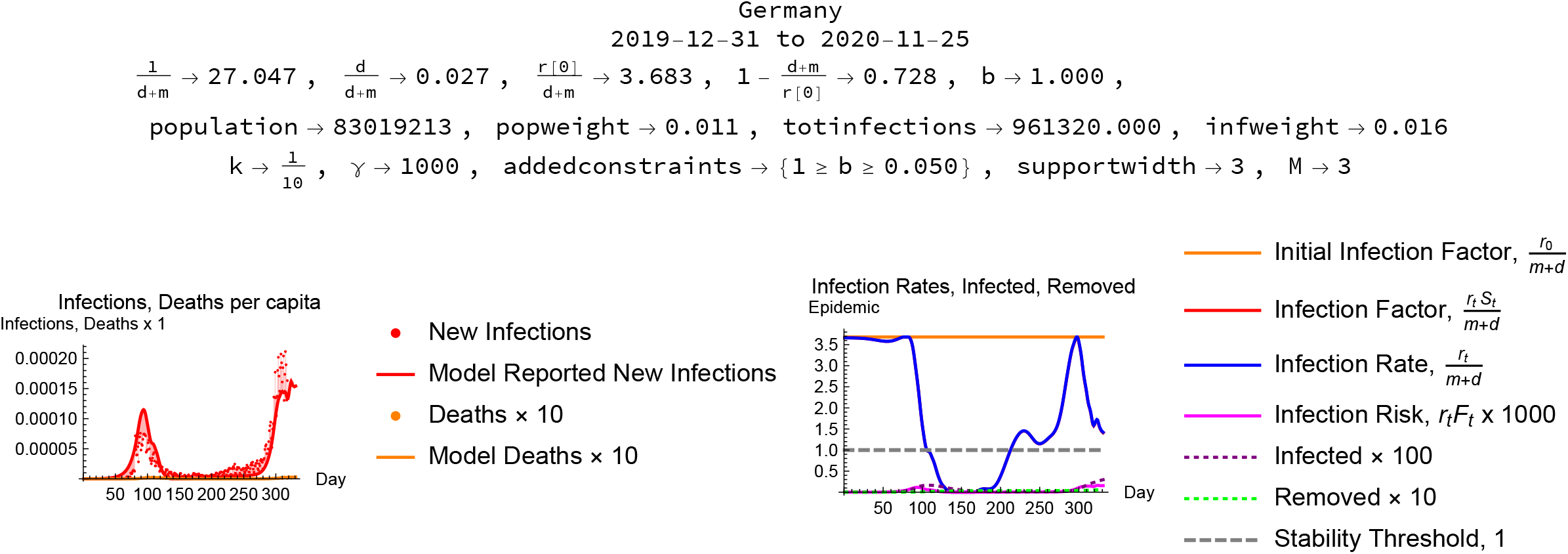
Germany.

### United Kingdom

**Figure 8:**
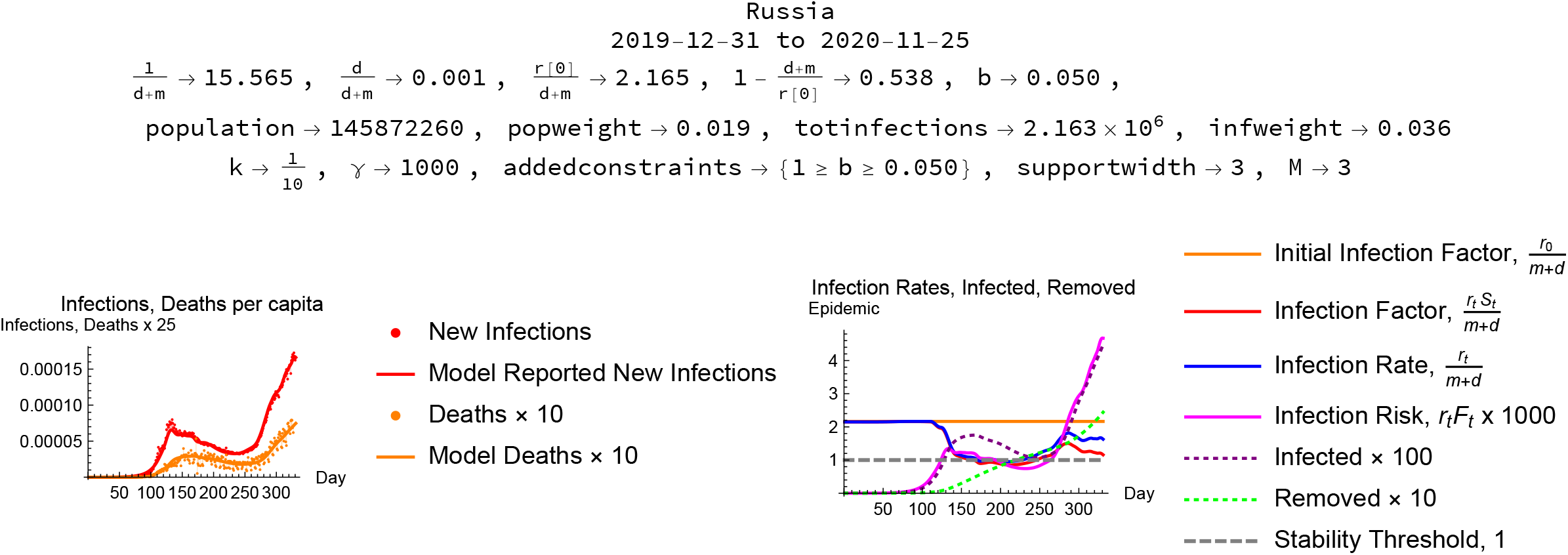
Russia.

### Mexico

**Figure 9:**
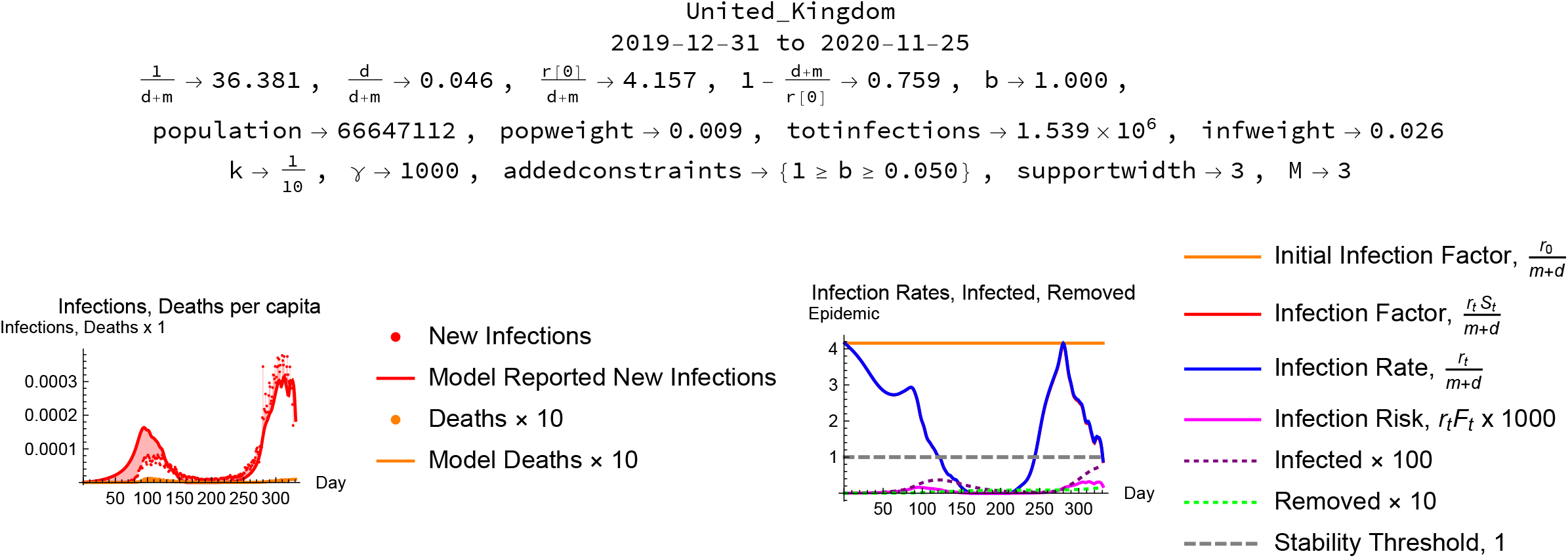
United Kingdom.

### Sweden

**Figure 10:**
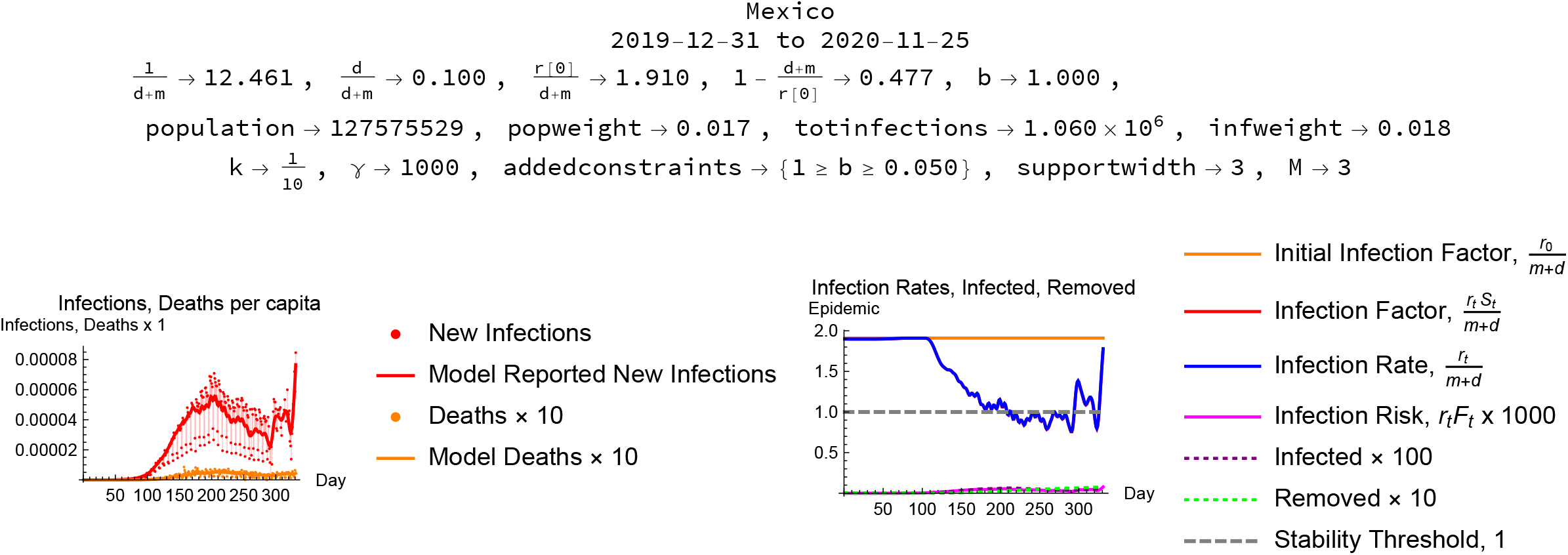
Mexico.

**Figure 11:**
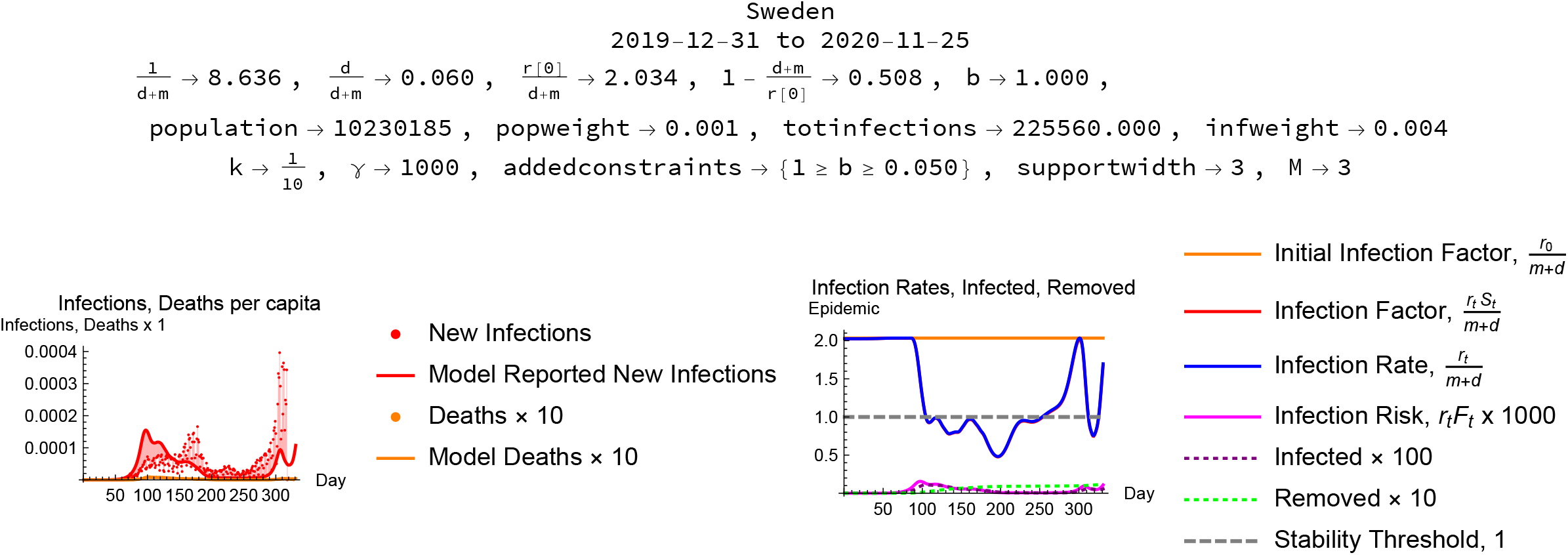
Sweden.

### Anomalous cases

Some country epidemic data result in very short (less than one day) or very long (over fifty days) infection lengths even with a constraint on the reporting ratio. An example is presented in Figure 12.

**Figure 12:**
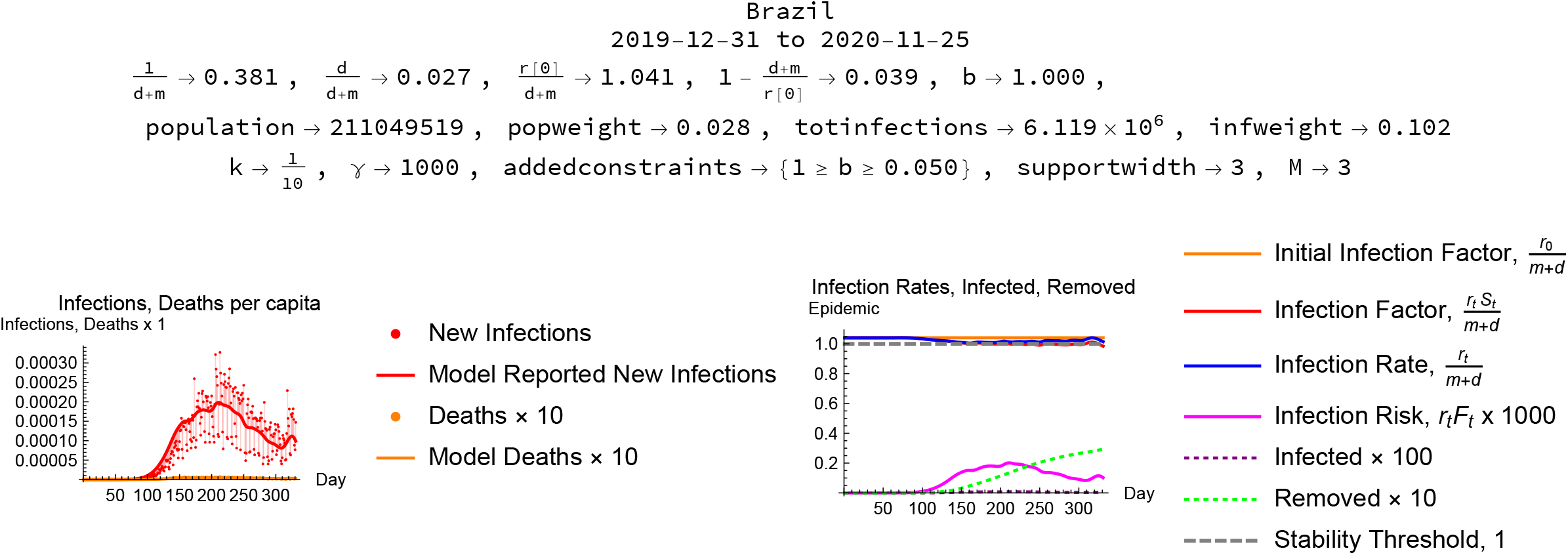
Brazil.

**Figure 13:**
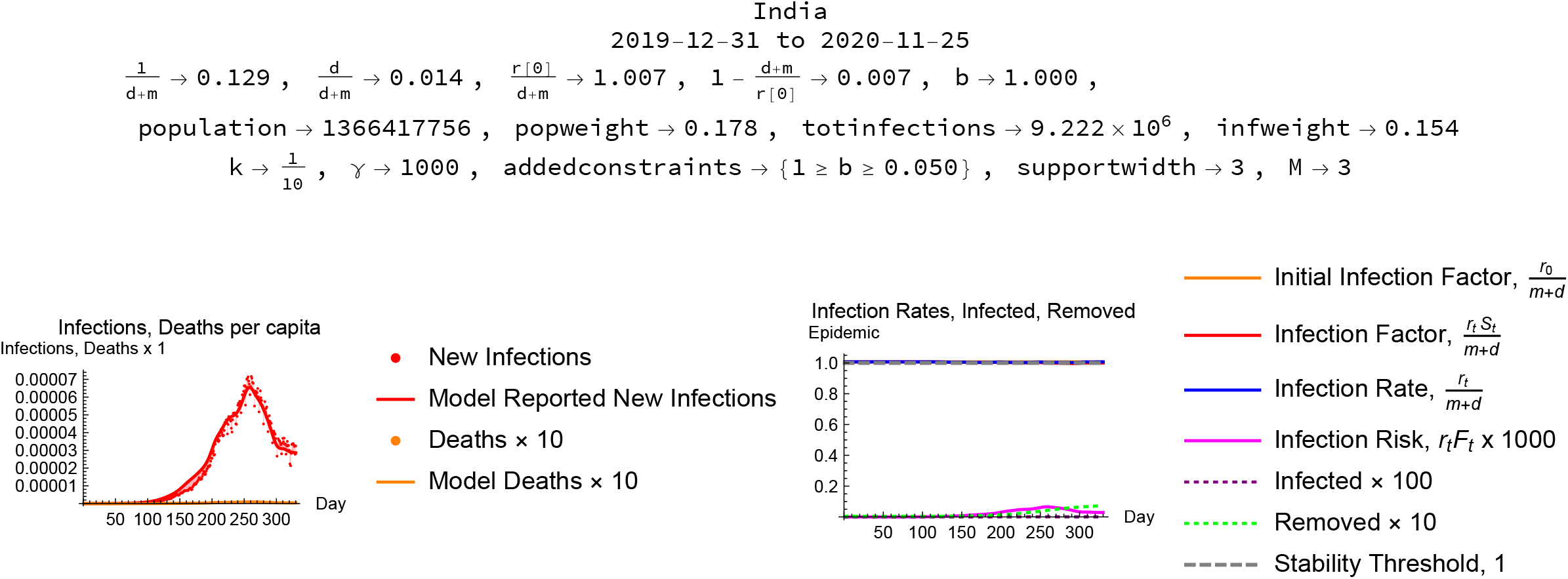
India.

### India

The data from India cannot be modeled successfully with the infection length constraint. The Indian case modeled without constraints is presented in Figure

## Supporting information

Updated Mathematica notebook for CME modeling

## Data Availability

Data is publicly available from sources cited in the paper.

https://www1.nyc.gov/site/doh/covid/covid-19-data.page

https://www.ecdc.europa.eu/en/publications-data/download-todays-data-geographic-distribution-covid-19-cases-worldwide

## Conclusions – modeling

The results of this modeling effort point to some conclusions both at the level of method and at the level of understanding the social aspects of an epidemic.

The model uses limited and noisy data, and reflects both the limitations of conclusions that can be drawn from the data and the potential of the info-metric constrained maximum entropy methods to extract whatever information a data set may contain. It is particularly striking that in many cases the constrained maximum entropy model allows us to draw fairly strong inferences about the behavior of unobserved, or not directly observed, variables like the daily infection factor without making strong assumptions such as linearity. There are undoubtedly numerous other ways to approach these same data sets with the CME methods, so it would be a mistake to argue that any particular CME model exhausts the information in the data, but given the constraints constituting any particular model the CME approach does promise to extract all the information available in that framework.

### Conclusions – epidemiology

The epidemiological results reflect limits not only of the quality and extent of the data but also of the model constraints. The greatest uncertainties arise because estimates of reporting rates, base infection factors and the length of the infectious period rest on extrapolation from limited observations of the initial stages of each epidemic and, as a result, are themselves fairly noisy. The results constitute a strong case that Covid-19, which has a half-life of infectiousness measured in a few days and is infectious enough to create an epidemic, is extremely sensitive to changes in social behavior. Whether epidemics have been controlled primarily by changes in social behavior or by exhaustion of the pool of susceptible individuals depends on uncertain estimates of the reporting rate. The simulations strongly support the view that measures that reduce infectiousness below the stability threshold can stabilize the epidemic in a relatively short time due to social multiplier effects. Relaxation of social distancing allows the infection factor to rise above unity and leads to a resurgence of the infection.

### Covid-19

Covid-19 is a particularly intractable phenomenon precisely because it is not very infective but it is quite lethal. Its infection rate is not high enough to establish herd immunity without tolerating loss of life on a scale unacceptable to many societies. Fortunately an understanding of the social multiplier dynamics governing the infection show that relatively low-cost measures to reduce the infection factor can be remarkably effective in suppressing the disease.

## Footnotes

2 A Mathematica notebook containing the programs used to generate these results is available for download at https://www.dropbox.com/s/8oe9avsyurq485x/EpidemicCMEModel20201127.nb?dl=0. The notebook is also available as supplementary material at the medRxiv server.

## References

Josh Epstein. Generative Social Science: Essays in Agent-based Computational Modeling. Princeton University Press, 2006.

Amos Golan. Foundations of Info-Metrics. Modeling and Inference with Imperfect Information. Oxford University Press, 2017.

David Smith and Lang Moore. The SIR model for spread of disease - the differential equation model. Convergence, 2004.

